# Obesity in the GLP-1 Era: Clinical Characteristics and Prescribing Patterns in a Large Integrated Health System

**DOI:** 10.64898/2026.09.04.26361064

**Authors:** Brian J Wells, Kristin M Lenoir, Michael Webster-Clark, Nicholas M Pajewski, Amanda T Pons-Junkins, Michelle M Mielke

## Abstract

**Aims:** To characterize adults with obesity in a large integrated health system.

**Methods:** We created a cross-sectional cohort of adults with at least one body mass index (BMI) value >=30 kg/m2 in the prior 18 months, a recent in-person outpatient visit, and an assigned primary care provider. Electronic health record data were transformed into a patient-level analytic dataset and linked to neighborhood-level social determinants of health. Diagnoses, laboratory values, vital signs, and prescriptions were curated using reproducible phenotype definitions and internally reviewed vocabularies.

**Results:** The registry included 280,186 adults, of whom 48% had class I obesity. The cohort was 63% female, 27% Black, and 14% lived in areas with high or very high deprivation. Common comorbidities included hypertension, type 2 diabetes, depression, and anxiety. HbA1c and cholesterol measurements were absent in more than 40% of the cohort during the prior two years. Overall, 22% of patients received a prescription for at least one weight loss drug in the past 2 years, including 18% who received a weight-loss-approved GLP-1 receptor agonist.

**Conclusions:** This care-engaged obesity cohort was characterized by high representation of women and socially vulnerable populations, substantial cardiometabolic screening gaps, and broad adoption of GLP-1 agonists.

## Introduction

The United States has one of the highest rates of obesity worldwide.(Nguyen et al., 2008) Obesity is a complex chronic disease associated with hypertension, type 2 diabetes, cardiovascular disease, chronic kidney disease, depression, and numerous other conditions. Understanding the epidemiology and management of obesity requires high-quality real-world data that can inform prevention strategies, guide clinical interventions, and monitor rapidly evolving patterns of treatment.

Large national datasets have provided important insights into the burden of obesity and its associated conditions. An analysis of administrative insurance claims data among middle-aged adults demonstrated that the prevalence of hypertension and diabetes was approximately two- and three-fold higher, respectively, among individuals with obesity compared to individuals who are normal weight.(Bae et al., 2025) However, claims-based datasets are generally limited by an absence of direct anthropometric measurements such as body mass index (BMI), prescriptions written, problem list diagnoses, and limited information on social determinants of health (SDoH). Alternatively, the National Health and Nutrition Examination Survey (NHANES) provides nationally representative estimates of obesity prevalence based on measured height and weight and includes SdoH (Li et al., 2022), but many diagnoses and medication data in NHANES rely on participant self-report or short-term recall. In addition, public data releases lag rapidly evolving clinical practice (for example, Medicare data is currently only available through 2022). Health system-derived electronic health record (EHR) data can complement these sources by providing a timely view of clinical care delivery, including vital signs, laboratory testing, diagnoses, prescribing behavior, and patient context. This capability is especially important in the context of obesity, where the therapeutic landscape is rapidly evolving.

Phentermine was approved for weight loss in 1959 and was followed by several appetite suppressants intended primarily for short-term use.(Tchang et al., 2024) Longer-term pharmacologic options emerged later, including orlistat, phentermine-topiramate, naltrexone-bupropion, device-based approaches such as Plenity, and lorcaserin, which has since been removed from the market. More recently, glucagon-like peptide-1 receptor agonists (GLP-1 receptor agonists) and related incretin-based therapies have transformed obesity treatment. Liraglutide was approved for chronic weight management in 2014, followed by semaglutide in 2021, tirzepatide in 2023, and an oral semaglutide formulation for weight loss in 2025.(Joszt, 2025; Tchang et al., 2024) Throughout this pharmacologic evolution, bariatric surgery has remained the most efficacious treatment for severe obesity and continues to play an important role alongside pharmacotherapy across the obesity treatment continuum.(Arterburn et al., 2020)

Pharmacy dispensing and prescription claims data have documented major shifts in obesity pharmacotherapy. Studies using Optum Clinformatics and IBM MarketScan data showed that phentermine-containing medications were the most commonly dispensed weight-loss drugs in 2018 (Suissa et al., 2021), followed by a transition toward GLP-1 receptor agonists as the dominant class by 2021-2022.(Ostrominski et al., 2025) EHR vendor-based federated real-world datasets may offer additional advantages. The value of Epic Cosmos data for studying GLP-1 receptor agonist use for pediatric weight management has also been demonstrated.(Mak et al., 2025) Because Epic-based federated datasets are derived from a shared EHR platform, they may offer greater structural consistency than distributed research networks (DRNs) that integrate heterogeneous source systems. However, these datasets do not replace locally validated, patient-level registries that can support phenotype curation, quality improvement, pragmatic trial recruitment, and analyses grounded in local clinical context. Other proprietary DRN platforms have provided important insights but also illustrate the need for complementary health system-based cohorts. For example, Rodriguez et al. used Truveta data (Rodriguez et al., 2025) to examine initiation and discontinuation patterns of GLP-1 receptor agonists and found substantial off-label use; approximately 62% of overweight or obese individuals without diabetes who initiated a GLP-1 receptor agonist received a diabetes-approved formulation. This finding underscores the need for medication groupings that can flexibly distinguish brand, ingredient, indication, and formulation. At the same time, the study focused on patients initiating GLP-1 therapy and did not evaluate the broader landscape of obesity pharmacotherapy or incorporate health system-specific care delivery patterns. More broadly, proprietary DRNs may combine prescriptions, dispensing, claims, EHR data, and SDoH information, but their de-identified structure limits direct utility for population health management, quality improvement, and clinical trial recruitment. Their development also requires extensive harmonization and local data quality assurance, processes that are not always transparent to end users and are difficult to assess centrally across participating sites.(Juárez et al., 2019; Kahn et al., 2015)

In response to these limitations, health systems can develop internal registries that allow greater flexibility in defining clinical phenotypes, validating data elements, supporting specific research objectives, and linking care delivery data to actionable quality improvement workflows. Real-world obesity registries provide an opportunity to characterize the population of patients with obesity who are actively engaged in clinical care and to examine how treatment patterns, comorbidity burden, and chronic disease management vary across demographic and social contexts. When linked to geocoded SDoH measures, such registries can support analyses of healthcare disparities, equitable care delivery, and targeted population health interventions.

A well-defined obesity cohort can also streamline pragmatic clinical trial recruitment embedded within these internal registries and reduce redundant data cleaning through validated phenotype definitions and standardized analytic variables. The purpose of this project was therefore to create a well-curated, cross-sectional, care-engaged cohort of adults with obesity in Atrium Health, the Southeast market of Advocate Health, together with an analytic dataset and accompanying variable definitions to support immediate quality improvement and research applications.

## Subjects, Materials, and Methods

### Registry Development and Data Source

The registry was developed by the Real-world Advocate DAta for Research (RADAR) team within the Clinical Trials Methods Center at Wake Forest University School of Medicine. RADAR includes professionals with expertise in statistics, epidemiology, informatics, programming, and clinical care and represents a collaborative partnership across the Departments of Epidemiology and Prevention, Biostatistics and Data Science, and the Clinical and Translational Science Institute. The team has experience extracting, cleaning, harmonizing, quality-checking, and analyzing EHR data for research and quality improvement. Registry development was informed by established principles for EHR data quality assessment, harmonization, and the secondary use of clinical data.(Kahn et al., 2012; Kahn et al., 2016)

Data were extracted from Atrium Health’s instance of the Epic EHR database (Clarity) using Microsoft SQL Server. This Epic implementation includes data across the Atrium Health network of 40 hospitals and approximately 1,400 care locations in the southeastern United States, including North Carolina, South Carolina, Georgia, and Alabama. Extracted data were transformed using R (version 4.5.1) from normalized relational tables into a patient-level analytic dataset. Data were extracted for two years before the index date of November 1, 2025.

### Cohort Definition

Inclusion criteria were:

- At least one instance of BMI ≥ 30 in the previous 18 months
- 1 outpatient, in-person visit in the 2 years prior to the index date.
- Atrium affiliated primary care provider noted in the associated EHR field on the index date.
- Age between 18 to 100 on the index date Exclusion criteria were:
- Prenatal visit or pregnancy-related ICD-10 codes within the year prior to the index date.
- Receiving hospice or palliative care
- Diagnosis of dementia or mild cognitive impairment in the 2 years prior to the index date
- Receiving dialysis
- eGFR <30 mL/min indicating both acute or longer-term kidney function issues
- Address in a state outside of the catchment area (NC, VA, SC, TN, WV, GA, AL).

Figure 1 provides a study flow chart to visualize the impact of the inclusion and exclusion criteria.

**Figure 1.**
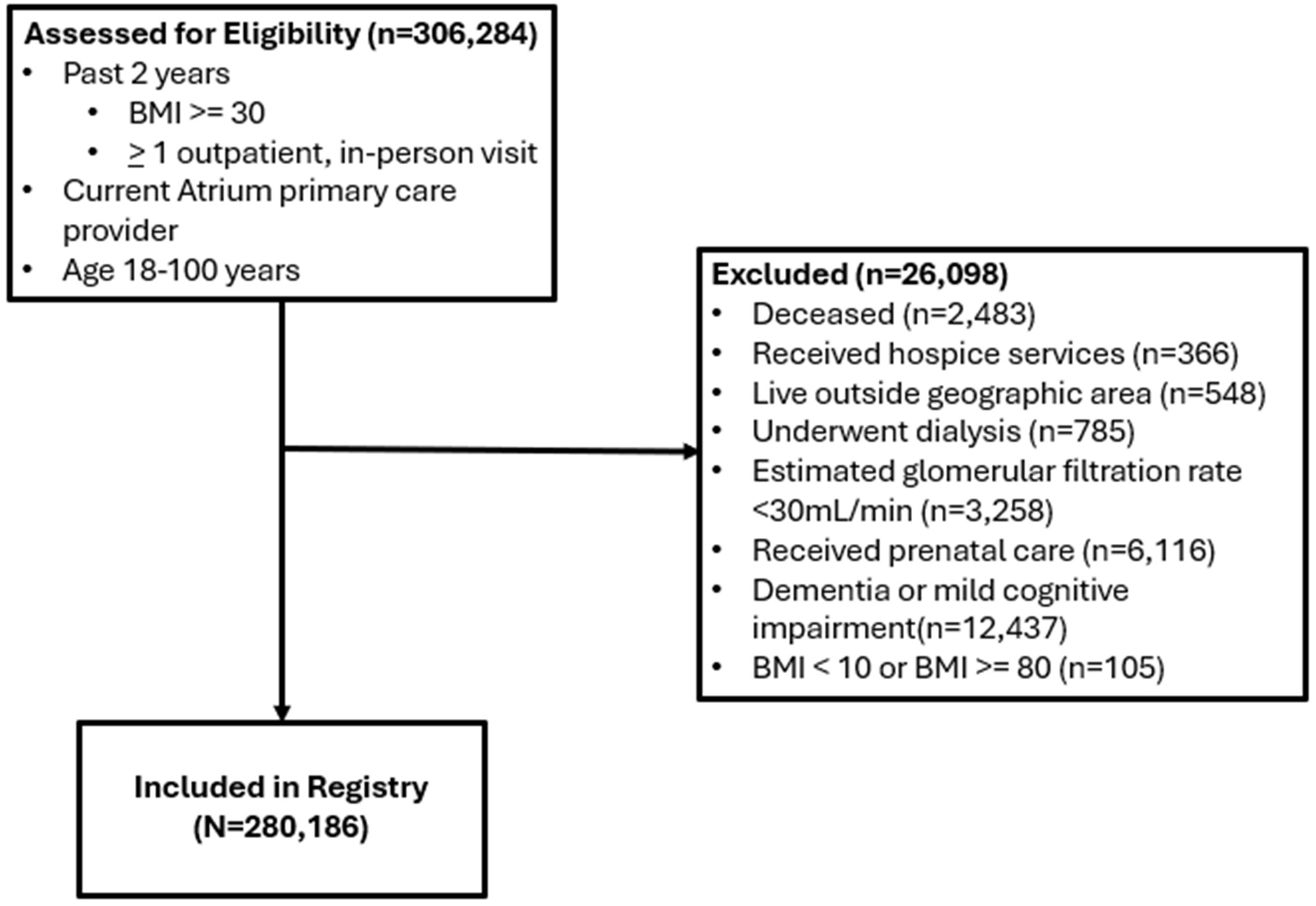
Study Flowchart

### Diagnoses

ICD-10 codes from structured fields in the EHR were grouped into comorbid conditions based on published groupings used in the Elixhauser comorbidity index.(Quan et al., 2011) Comorbidities were identified from encounter and active problem list diagnoses during the two-year lookback period; a single qualifying diagnosis was sufficient for classification. The two-year lookback period was selected to emphasize clinically active or recently documented conditions and to characterize the current health status of patients actively receiving care within the health system. Curated diagnosis code lists are provided in the supplementary materials. The electronic frailty index (eFI) was limited to patients at least 55 years of age who had two outpatient visits in the prior two years and sufficient completed data elements, consistent with the original method.(Pajewski et al., 2019)

### Laboratory Values

The team selected a limited list of laboratory values particularly relevant to patients with obesity including glucose, hemoglobin A1c, cholesterol, liver enzymes, and kidney function. Kidney function was quantified using estimated glomerular filtration rate (eGFR) calculated from serum creatinine using formulas that do not include race as a predictor.(Inker Lesley A. et al., 2021) Laboratory values in the analytic dataset represent the most recent available result on or before November 1^st^, 2025. Glucose, triglycerides, liver enzymes, and creatinine values obtained in emergency department, inpatient, or observation settings were excluded because they are sensitive to acute illness.

Because key identifiers such as LOINC codes, specimen type, units, and reference ranges were inconsistently populated, laboratory classification required a multi-attribute approach using available metadata fields. The team established biologically plausible ranges for each laboratory test using clinical expertise and published literature. Laboratory records outside these ranges were removed, and the next available valid result for the patient was used.

### Medication Classification

Prior studies have shown that manual curation and validation of medication code lists may improve classification accuracy and reproducibility compared with reliance on standard classification systems alone. (McDonough et al., 2020; Graul et al., 2023) Medication classification was therefore based on an internally developed vocabulary created by the RADAR team. Medication lists were compiled using multiple authoritative sources, including the Anatomical Therapeutic Chemical classification system (World Health Organization, 2025), RxNorm (National Library of Medicine, 2025), First Databank (First Databank, 2025), and Epocrates (Epocrates, 2025). Each entry was reviewed and verified by at least two team members. After compiling medication names, the team used text-string searching similar to the approach described by Graul et al. (Graul et al., 2023). The approach identifies and assigns combination medications such as phentermine-topiramate to multiple classes while permitting separate assessment of brand-specific and ingredient-based GLP-1 receptor agonist categories. The registry was limited to outpatient oral, transdermal, and subcutaneous prescriptions to capture formulations consistent with longitudinal outpatient management and excluding formulations unlikely to represent the intended indication. All brand and generic agents included in each class are provided in the accompanying reference materials. Medication exposure was defined as one or more prescription orders for a drug within a given class during the two years prior to November 1, 2025.

### Vital Signs, Demographics, Social History and SDoH

Baseline height was defined as the median or modal biologically plausible height value recorded anywhere in the EHR on or after the patient’s 18th birthday. Other vital signs were defined using the most recent value on or before November 1, 2025. If multiple blood pressure values were available during the same encounter or on the same date, the most recent value was used. Weights and blood pressure obtained during inpatient, emergency department, or observation encounters were excluded. Race and ethnicity categories were coded to align with current NIH guidelines. Age was calculated on the index date. Because smoking history was inconsistently documented in structured fields, pack-years, vaping history, and cigar use were not calculated. Smoking status was defined using the most recent social history entry with a known status. Patient addresses were geocoded at the block group level and linked to American Community Survey data to obtain census-derived SDoH measures.(US Census Bureau, 2025) These data were used to obtain Area Deprivation Index national percentile rankings, which were dichotomized into high/very high deprivation (>=85^th^ percentile) versus lower deprivation (<85^th^).(Flanagan et al., 2011; Kind and Buckingham, 2018) Urban-rural status was assigned using residential ZIP codes linked to the 2020 Rural-Urban Commuting Area classification.(Economic Research Service, 2026)

## Results

There were 306,284 potentially eligible patients in the Atrium Health system with BMI >=30 kg/m2 with an Atrium-affiliated primary care provider, and at least one in-person outpatient visit in the 18 months prior to November 1, 2025. Exclusions included 6,116 women who received prenatal care in the prior year, 12,437 patients with dementia or mild cognitive impairment, and 3,258 patients with eGFR <30 mL/min. Table 1 summarizes descriptive characteristics for the 280,186 patients meeting all eligibility criteria. Approximately 48% of patients were classified as obesity class I (BMI 30.0-34.9 kg/m2), 26% as class II (BMI 35.0-39.9 kg/m2), and 22% as class III (BMI greater >= 40 kg/m2). Women represented 63% of the cohort and accounted for 72% of patients with class III obesity. Black patients represented 27% of the overall cohort and 34% of patients with class III obesity. Most patients resided in North Carolina (81%), 14% lived in census block groups with high-very high levels of deprivation, and 11% had Medicaid listed as one of their insurance providers on the index date.

**Table 1.**
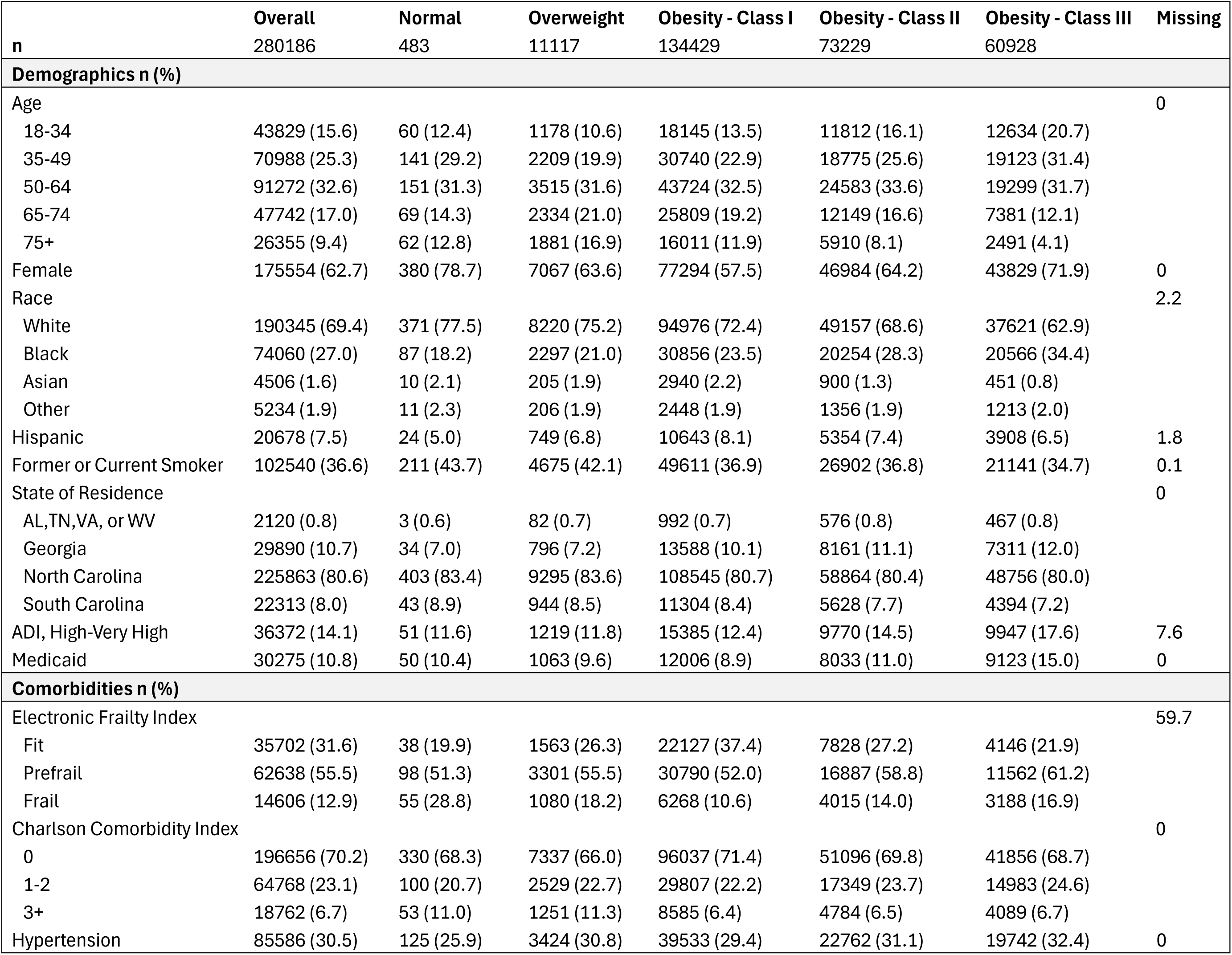
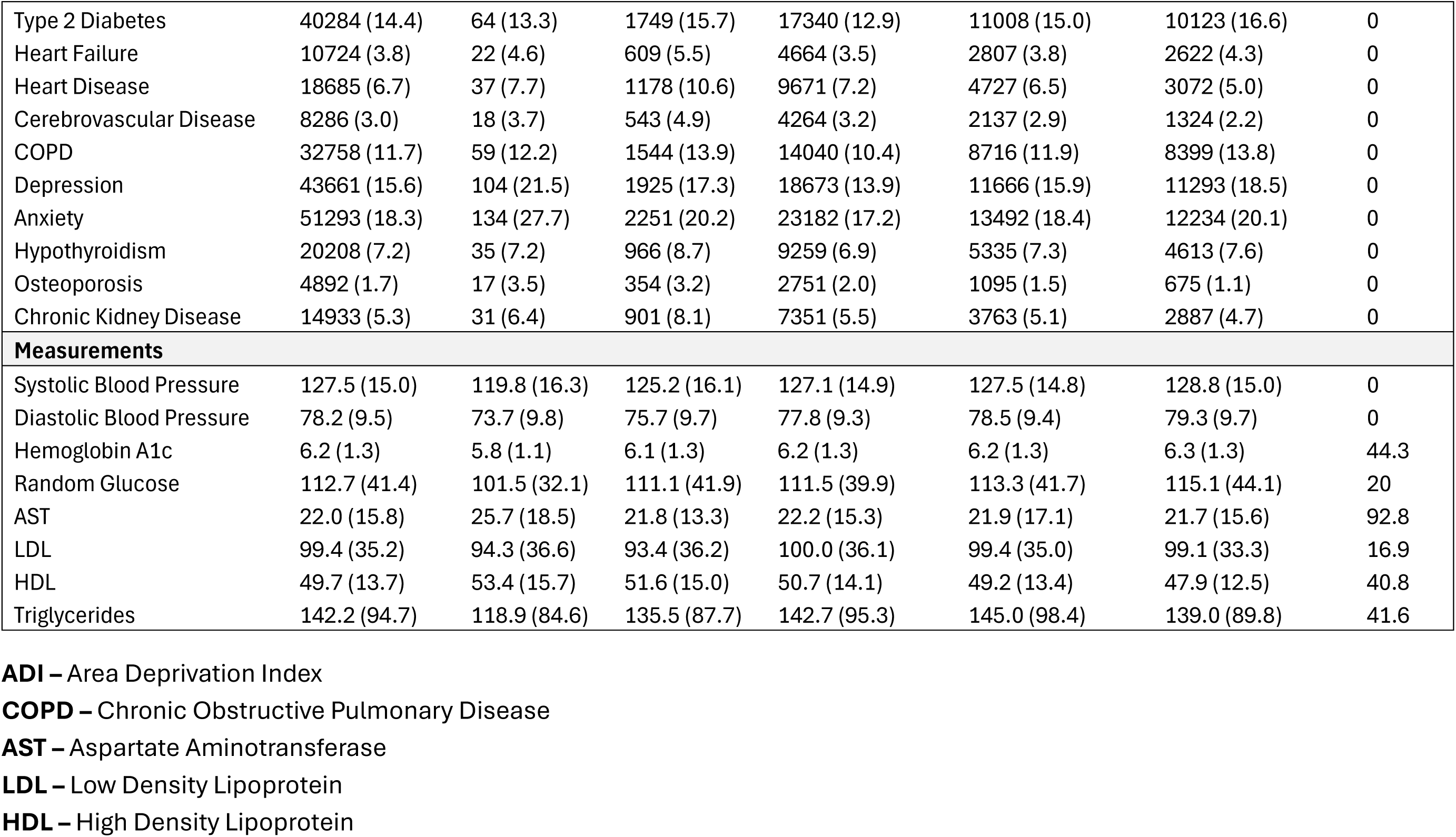
Characteristics of Patients with Obesity at Atrium Health by Current Weight Status.

The most common comorbidities were hypertension (31%), anxiety (18%), and depression (16%). Increasing obesity class was associated with higher mean systolic and diastolic blood pressure, random glucose, and hemoglobin A1c (HbA1c). HbA1c and cholesterol measurements were missing in 44% and 41% of the cohort, respectively, during the prior two years. A type 2 diabetes diagnosis in the prior two years was present in 14% of the cohort. Prescription patterns differed substantially by diabetes status (Table 2). Antihypertensive prescriptions were more common among patients with type 2 diabetes than among those without (85% vs. 57%), as were prescriptions for glucose-lowering medications (88% vs. 38%). In contrast, weight-loss-specific medications were more common among patients without type 2 diabetes (24% vs. 9%). GLP-1 receptor agonists approved for weight management (Wegovy, Saxenda, and Zepbound) were the most commonly prescribed weight-loss medication category, with 18% of patients receiving at least one prescription during the prior two years.

**Table 2.** Prescriptions in the Previous 2 Years by Type 2 Diabetes Status.

|  | Overall | FALSE | TRUE |
| --- | --- | --- | --- |
| n | 280186 | 239902 | 40284 |
| **Any Antihypertensive** | **171693 (61.3)** | **137608 (57.4)** | **34085 (84.6)** |
| Ace Inhibitors | 56659 (20.2) | 42297 (17.6) | 14362 (35.7) |
| Alpha Agonists | 5716 (2.0) | 4612 (1.9) | 1104 (2.7) |
| ARBs | 69856 (24.9) | 54799 (22.8) | 15057 (37.4) |
| Beta-blockers | 70413 (25.1) | 55201 (23.0) | 15212 (37.8) |
| Calcium Channel Blockers | 65105 (23.2) | 52089 (21.7) | 13016 (32.3) |
| Potassium Sparing Diuretics | 20574 (7.3) | 16176 (6.7) | 4398 (10.9) |
| Thiazide Diuretics | 66337 (23.7) | 52728 (22.0) | 13609 (33.8) |
| Vasodilators | 5745 (2.1) | 4119 (1.7) | 1626 (4.0) |
| **Any Glucose Lowering Medication** | **126231 (45.1)** | **90629 (37.8)** | **35602 (88.4)** |
| Alpha Glucosidase Inhibitors | 195 (0.1) | 121 (0.1) | 74 (0.2) |
| Amylin Mimetics | 6 (0.0) | 2 (0.0) | 4 (0.0) |
| Metformin | 56933 (20.3) | 32602 (13.6) | 24331 (60.4) |
| DPP4 Inhibitors | 5868 (2.1) | 2903 (1.2) | 2965 (7.4) |
| GLP-1 Agonists | 97276 (34.7) | 73083 (30.5) | 24193 (60.1) |
| Insulin | 1376 (0.5) | 731 (0.3) | 645 (1.6) |
| Meglitinides | 137 (0.0) | 66 (0.0) | 71 (0.2) |
| SGLT2 Inhibitors | 22933 (8.2) | 12473 (5.2) | 10460 (26.0) |
| Sulfonylureas | 15258 (5.4) | 7431 (3.1) | 7827 (19.4) |
| Thiazolidinediones | 4943 (1.8) | 2402 (1.0) | 2541 (6.3) |
| **Any Weight Loss Drug** | **60403 (21.6)** | **56895 (23.7)** | **3508 (8.7)** |
| Weight Loss Approved GLP-1 | 50531 (18.0) | 47884 (20.0) | 2647 (6.6) |
| Wegovy | 34386 (12.3) | 32689 (13.6) | 1697 (4.2) |
| Zepbound | 28604 (10.2) | 27294 (11.4) | 1310 (3.3) |
| Saxenda | 2669 (1.0) | 2523 (1.1) | 146 (0.4) |
| Non GLP-1 Agonist | 20347 (7.3) | 19119 (8.0) | 1228 (3.0) |
| Contrave | 1958 (0.7) | 1839 (0.8) | 119 (0.3) |
| Qsymia | 1276 (0.5) | 1220 (0.5) | 56 (0.1) |
| Plenity | 31 (0.0) | 29 (0.0) | 2 (0.0) |
| Stimulant | 17948 (6.4) | 16872 (7.0) | 1076 (2.7) |
| Orlistat | 272 (0.1) | 252 (0.1) | 20 (0.0) |

Table 3 shows weight-loss drug prescribing patterns across demographic and social groups. Overall, 22% of patients received at least one weight-loss drug prescription, 18% received a weight-loss-approved GLP-1 receptor agonist, and 6% received a stimulant medication. Weight-loss-approved GLP-1 receptor agonists were the most commonly prescribed class across sex, age, race, ADI category, and Medicaid status. Women in the cohort were more than twice as likely as men to receive any weight-loss drug prescription (27% vs. 13%), with prescribing peaking at 36% among women aged 35-49 years. Prescribing was less common in older adults; only 5% of patients aged 75 years or older received at least one weight-loss drug prescription. Non-GLP-1 weight-loss medications were uncommon, with Contrave the most common but only prescribed to 0.7% of the cohort.

**Table 3.**
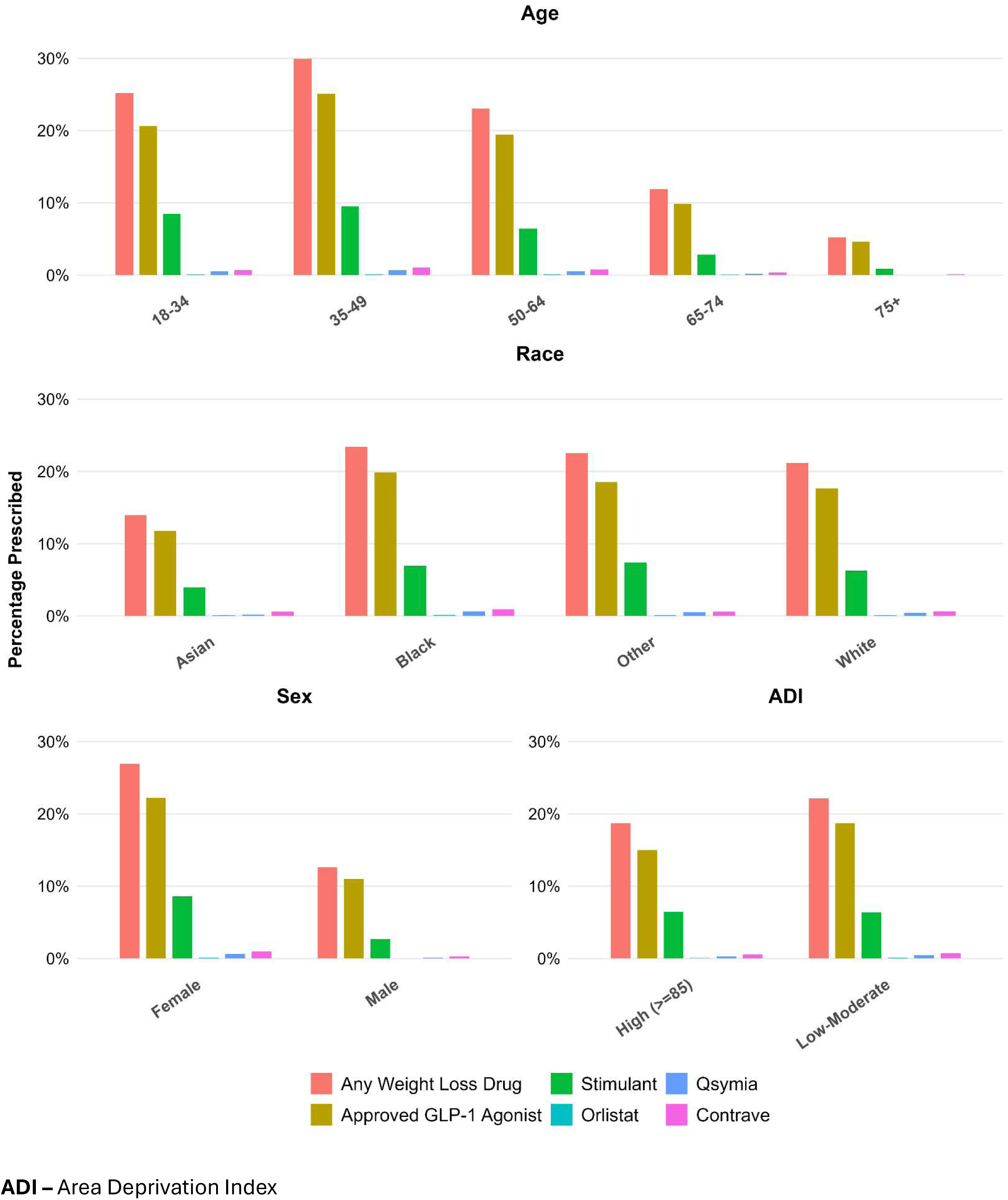
Weight Loss Drug Prescriptions by Demographics.

## Discussion

This registry provides a care-delivery benchmark for obesity management at scale in a large integrated health system. Rather than representing only patients seen in specialty weight-management clinics or patients that are currently filling prescriptions for anti-obesity medications, the cohort captures adults with obesity who were actively engaged in routine primary care. The registry therefore describes what obesity-related comorbidity assessment and pharmacotherapy prescribing look like in real-world practice across a large health system serving a racially and socially diverse population. The demographic composition of the cohort differed from the U.S. population in important ways. Black patients represented 27% of the cohort and women represented 63%, compared with approximately 12.5% and 51% of the U.S. population, respectively.(Dimitry, 2025) The high representation of Black patients and individuals living in areas with high deprivation creates an important opportunity to evaluate and address obesity-related health disparities. Women accounted for 72% of patients with class III obesity, a proportion higher than that reported among patients with comparable BMI ranges. (MacEwan et al., 2024)

The prevalence of type 2 diabetes in our cohort increased from 12.9% in class I to 16.6% in class III obesity, consistent with the graded relationship reported by Nguyen et al. using NHANES data. (Nguyen et al., 2008) However, the prevalence in that study was only 14.2% among patients with class III obesity. Hypertension prevalence showed a more modest gradient across obesity classes (29.4% in class I to 32.4% in class III) compared with the steeper increase observed in NHANES (34.0% to 52.3%), (Nguyen et al., 2008) which may reflect differences in ascertainment methods; NHANES defines hypertension by measured blood pressure and medication use, whereas our registry relies on ICD-coded diagnoses. Differences between our findings and those reported in NHANES may also reflect variation in geography, data sources, cohort eligibility criteria, exclusion criteria, and comorbidity definitions.

The cohort appeared relatively healthy by Charlson comorbidity count, with 70% of patients having no Charlson-defined condition. However, the Charlson index does not include common obesity-related conditions such as hypertension, and the registry includes a care-engaged outpatient population subject to planned exclusions that favor healthier individuals. Depression and anxiety were common. Given the well-established bidirectional relationship between depression and obesity (Milaneschi et al., 2019) the registry will support future longitudinal analyses of whether obesity treatment is associated with changes in depression-related diagnoses, medication use, or healthcare utilization.

The registry also identified potential opportunities for cardiometabolic screening and risk management. Mean blood pressure, HbA1c, AST, and lipid values generally increased with obesity class, and more than 40% of patients lacked HbA1c or lipid measurements in the prior two years. These findings suggest potential gaps in screening, although interpretation requires caution because individualized risk profiles, prior testing outside the health system, and clinical indications for testing were not assessed in this cross-sectional analysis.

Our findings are consistent with prior work showing that GLP-1 agonists have become the dominant class of weight-loss pharmacotherapy.(Ostrominski et al., 2025) In this registry, weight-loss-approved GLP-1 agonists were the most commonly prescribed weight-loss medication category across all major demographic strata. However, prescription records represent clinician intent and patient access efforts, not medication acquisition or use. Prior research suggests that many GLP-1 receptor agonist prescriptions for weight loss go unfilled, likely due to cost, insurance coverage, and medication availability.(Gilbert et al., 2025) Among patients with obesity and without type 2 diabetes, only 37% filled a GLP-1 receptor agonist prescription despite continuous insurance enrollment.(Sarpatwari et al., 2025)

Temporal factors further complicate interpretation of obesity pharmacotherapy. Wegovy, Zepbound, and Saxenda were each listed by the FDA as being in shortage at least once between 2022 and 2025, during which compounded formulations were permitted under certain circumstances despite not being evaluated by the FDA for safety, quality, or effectiveness. These formulations may not be captured in the EHR when obtained through direct-to-consumer or telehealth-based platforms. These issues highlight the importance of longitudinal studies designed to examine initiation, discontinuation, switching, and reinitiation over time.

Beyond its immediate value for describing obesity care, the registry developed for this study will create a foundation for future pharmacoepidemiologic investigations of the broader systemic effects of weight-loss therapies, including long-term outcomes not fully assessed in initial randomized trials. For example, GLP-1 receptor agonists have been hypothesized to confer risks and benefits beyond weight loss and glycemic control, with growing interest in their potential effects on addictive behaviors; recent studies suggest they may reduce cravings not only for food, but also for alcohol and other substances.(Hendershot et al., 2025) A curated, health system-based registry with validated phenotypes and the ability to link to longitudinal outcomes can support future analyses of these questions while also facilitating pragmatic trial recruitment and targeted quality improvement.

### Limitations and Future Directions

Registries such as the one presented in this paper create opportunities for pragmatic clinical trials involving patients with obesity. These include studies that leverage EHR infrastructure for trial conduct—such as feasibility assessment, study recruitment, data collection, patient communication, and outcome ascertainment (Conroy et al., 2019; Thomas et al., 2024)—as well as those that incorporate interventions directly into the EHR (e.g., clinical decision support, alerts, order sets, documentation tools, and care pathways). To date, only a limited number of EHR-integrated intervention studies addressing obesity have been conducted, and most have involved sample sizes in the hundreds.(Ackermann et al., 2025; Baer et al., 2020; Conroy et al., 2019) One notable exception is the PATHWEIGH pragmatic trial, a stepped-wedge cluster-randomized trial conducted across 56 primary care clinics in Colorado that evaluated an EHR-driven care process consisting of leadership endorsement, an EHR-integrated weight management workflow, and clinician education and implementation strategies, with outcomes assessed among 274,182 patients.(Perreault et al., 2026) Our population may offer advantages in terms of racial and ethnic diversity; for example, Black patients comprised 4% of the PATHWEIGH analytic sample compared with 27% in our registry.

The registry also provides the opportunity to identify patients for pharmacologic pragmatic trials, such as the recently completed trial comparing semaglutide with other commercially available weight loss medications.(Peters et al., 2026) The large sample size within a single health system offers a substantial pool of potential participants, reducing the resources, complexity, and limitations associated with conducting multisite pragmatic clinical trials. The Trial Innovation Network (TIN) and Recruitment Innovation Center (RIC) have developed an EHR-based cohort assessment resource that provides summary counts of patients with conditions of interest across Clinical and Translational Science Awards (CTSA) hubs. However, because it is primarily a feasibility-counting tool, it does not directly determine how many patients would meet protocol-specific eligibility criteria, provide patient-level data, or create a mechanism for identifying and recruiting individual patients.(Nelson et al., 2022)

This registry should be viewed as an initial step toward the use of EHR data for real-world analyses of obesity. Its current design provides a clinically useful cross-sectional characterization of adults with obesity actively engaged in care, but several limitations should be acknowledged. Although the cohort includes a large number of patients across multiple states, it remains limited to one region of the United States. Planned expansion to Advocate Health’s Midwest region will improve geographic generalizability. The registry does not yet capture key elements of obesity management that would provide a more complete picture of care delivery, including bariatric surgery, referrals to obesity medicine or bariatric specialists, referrals to nutrition or diabetes education services, and the specialty or provider type of clinicians prescribing obesity-related therapies. These omissions currently limit the ability to evaluate treatment pathways across the full continuum of obesity care.

The current analyses are best interpreted as describing a care-engaged population of patients with obesity rather than all adults with obesity in the underlying service area. Because inclusion required recent contact with the health system, the registry may preferentially capture individuals with greater healthcare utilization, more documented comorbidity, or more opportunities for diagnosis and treatment. Sex differences in healthcare engagement may further shape cohort composition, as women are more likely than men to maintain regular contact with primary care, which may partly explain the higher representation of women. This design was appropriate for the present descriptive objectives, but alternative inclusion criteria may be preferable for future pharmacoepidemiologic or longitudinal investigations.

Fasting status was not reliably available; therefore, all glucose measurements were treated as random glucose values. This limitation may affect interpretation of glucose levels and comparisons across patients. Similarly, BMI-based classification has known limitations in certain clinical contexts. The registry does not yet account for factors that may distort BMI as a marker of adiposity, including edema related to heart failure and weight estimates in patients with limb amputations. In addition, the current version does not explicitly incorporate prior weight trajectories, so it may misclassify individuals whose recent BMI does not reflect substantial prior weight loss or weight gain. The registry currently excludes adults who are overweight but not obese. This group is important for prevention and early intervention because some patients with BMI 27 to <30 kg/m2 and obesity-related comorbidities, such as obstructive sleep apnea, may be candidates for pharmacologic treatment after unsuccessful lifestyle intervention.(AGA Clinical Guidelines Committee et al., 2022) Expanding the registry to include this population will enhance its value for research on prevention, early intervention, and obesity progression, but would require additional storage, computation, and data export capacity.

Our analysis was limited to the southeastern United States and excluded patients with dementia, dialysis, and eGFR <30 mL/min because one intended function of the registry is to support identification of patients potentially eligible for clinical trials. Our type 2 diabetes definition required a diagnosis code in the prior two years and did not incorporate HbA1c values, which likely improved contemporaneous specificity but may have reduced sensitivity. The true prevalence of diabetes in our health system is therefore likely higher than reported here.

Medication exposure is another important limitation. Outpatient prescriptions written within the health system do not confirm medication acquisition and administrative claims data fail to capture prescriptions that were paid for in cash, billed through insurance discount programs or through unregulated domestic or international channels. (Ashraf et al., 2024) Interpreting prescriber intent presents a further analytic challenge, particularly for medications with indications for multiple conditions. A clinician’s primary motivation is not readily discernible from the prescription record alone. Medications in this study were identified using both brand and generic names; however, future work should expand this framework to incorporate routes of administration, formulation strengths, and prescribed dosages. Such granularity would allow for a more nuanced understanding of prescriber intent, facilitate dose–response analyses, and improve comparability across treatment regimens.

Future work will focus on adding pharmacy dispensing information, incorporating time-varying longitudinal data, expanding clinical phenotype definitions, and linking to claims and additional outcomes. Using HIPAA-compliant large language model workflows to extract structured information from unstructured clinical text and other multimodal data may further enhance the registry. These steps will require careful validation, staged implementation, and continued quality assurance, but they will allow the registry to evolve from a cross-sectional care-delivery snapshot into a durable platform for obesity research, population health, and clinical trial recruitment.

## Conclusions

This EHR-based registry of over 280,000 adults with obesity engaged in primary care across a large, racially diverse health system reveals a graded increase in cardiometabolic comorbidity across obesity classes, substantial gaps in cardiometabolic screening, and the rapid emergence of GLP-1 receptor agonists as the dominant prescribed weight-loss pharmacotherapy, particularly among women aged 35 to 49 years. The scale, demographic diversity, and granular clinical and neighborhood-level data captured in this registry establish a robust platform for obesity-related health disparities research, pharmacoepidemiologic investigation, quality improvement, and pragmatic clinical trial recruitment. As obesity care continues to evolve with the expanding therapeutic landscape, registries of this kind will be essential for generating the real-world evidence needed to guide equitable, effective, and sustainable treatment at the population level.

## Supporting information

Supplemental material - medications and diagnoses codes

## Data Availability

All data produced in the present study are available upon reasonable request to the authors

## Acknowledgments

We would like to acknowledge the Informatics Program of the Wake Forest Clinical and Translational Science Institute (WF CTSI), which is supported by the National Center for Advancing Translational Sciences (NCATS), National Institutes of Health, through Grant Award Number UM1TR004929.

